# Localized and Widespread Chronic Pain in Sexual and Gender Minority People – An Analysis of The PRIDE Study

**DOI:** 10.1101/2023.11.27.23299101

**Authors:** Andrea L. Chadwick, Nadra E. Lisha, Micah E. Lubensky, Zubin Dastur, Mitchell R. Lunn, Juno Obedin-Maliver, Annesa Flentje

**Affiliations:** Department of Anesthesiology, Pain, and Perioperative Medicine, University of Kansas School of Medicine, Kansas City, Kansas; Division of General Internal Medicine, Department of Medicine, University of California, San Francisco, San Francisco, CA; Department of Community Health Systems, University of California, San Francisco, San Francisco, CA; The PRIDE Study/PRIDEnet, Stanford University School of Medicine, Stanford, CA; Department of Obstetrics and Gynecology, Stanford University School of Medicine, Stanford, CA; Division of Nephrology, Department of Medicine, Stanford University School of Medicine, Stanford, CA; Department of Epidemiology and Population Health, Stanford University School of Medicine, Stanford, CA; Alliance Health Project, Department of Psychiatry and Behavioral Sciences, University of California San Francisco, San Francisco, CA

## Abstract

Sex related differences, without taking gender into account, in chronic pain have been widely researched over the past few decades in predominantly cisgender and heterosexual populations. Historically, chronic pain conditions have a higher incidence and prevalence in cisgender women, including but not limited to fibromyalgia, irritable bowel syndrome, and migraine. The goal of the present study was to identify and characterize the presence and characteristics of chronic pain in SM and GM persons using data from The PRIDE Study, which is the first large-scale, long-term national cohort health study of people who identify as lesbian, gay, bisexual, transgender, queer, or as another sexual or gender minority person. A total of 6189 adult participants completed The PRIDE Study 2022 Annual Questionnaire at the time of data analysis. A total of 2462 participants reported no chronic pain, leaving 2935 participants who reported experiencing chronic pain. The findings from this study highlight that chronic pain is present to a significant degree in sexual and gender minority adults who participated in The PRIDE Study with chronic spine pain being the most common location/region of pain. Notably, more than one-third of non-binary persons, transgender men, and people who selected another gender experienced chronic widespread pain, defined by having 3 or more total regions of chronic pain. The lowest prevalence of chronic widespread pain was among transgender women and cisgender men. When considering sexual orientation, the highest prevalence of widespread pain was in participants who selected another sexual orientation, followed by queer and asexual, demisexual, gray ace, with the lowest prevalence of chronic widespread pain being in those who identify as straight or heterosexual, bisexual, pansexual, gay, and lesbian. Future studies are planned to elucidate how a variety of biopsychosocial mechanisms may influence chronic pain in sexual and gender minority persons.

## Introduction

Chronic pain is one of the most prevalent diseases worldwide and leads to substantial disability and enormous socioeconomic costs.^1^ Sex-related differences in chronic pain have been widely researched over the past few decades in predominantly cisgender (persons whose gender corresponds to their sex assigned at birth) and heterosexual populations. Historically, chronic pain conditions have a higher incidence and prevalence in cisgender women, including but not limited to fibromyalgia, irritable bowel syndrome, and migraine.^2^ Sexual minority (SM) persons (bisexual, gay, lesbian, or those with other non-heterosexual sexual orientations) and gender minority (GM) persons (those with a gender identity and/or expression that is different from what is traditionally expected based on the sex they were assigned at birth) comprise 7% and 0.6% of American adults, respectively.^3, 4^ As research in sexual and gender minority (SGM) persons has increased, data have suggested that higher rates of chronic pain syndromes are observed in SGM populations compared to heterosexual and cisgender populations.^3, 5^

The goal of the present study was to identify and characterize the presence and characteristics of chronic pain in SGM persons using data from The PRIDE Study, which is a large-scale, longitudinal, national cohort health study of people who identify as lesbian, gay, bisexual, transgender, queer, intersex, and asexual (LGBTQIA+), or as another sexual or gender minority person.

## Methods

Data were collected through The PRIDE Study.^6^ Inclusion criteria were (i) identifying as a SGM person, (ii) being at least 18 years old, (iii) living in the United States or its territories, and (iv) reading and understanding English. Data came from The PRIDE Study’s 2022 Annual Questionnaire and were collected between June 6, 2022 and May 16, 2023.^6^ Consent was obtained electronically, and institutional review board (IRB) approval was obtained from the University of California, San Francisco, Stanford University, and WIRB-Copernicus Group (WCG) IRBs. For our study, our primary outcome was the presence and location of chronic pain. To assess this outcome, participants were instructed to identify, on the Michigan Body Map,^7^ the regions of their body in which they have chronic pain or to report that they do not have chronic pain. We calculated the proportion of participants who reported chronic pain in seven body regions: A) left shoulder, left upper arm, left lower arm; B) right shoulder, right upper arm, right lower arm; C) left hip/left buttocks, left upper leg, left lower leg; D) right hip/right buttocks, right upper leg, right lower leg; E) neck, upper back, lower back; F) chest, abdomen; and G) right jaw and left jaw.^8^ We identified the proportion of participants who had chronic pain in greater than or equal to 3 regions of the body, which was defined as being positive for the presence of widespread pain.^8^

Participants in The PRIDE Study are asked to self-select their gender identity from the following list: cisgender men, cisgender women, non-binary, transgender men, transgender women, and another gender identity.^9^ They were also asked to self-select their sexual orientation from the following list: asexual/demisexual/gray-ace, bisexual/pansexual, gay/lesbian, heterosexual/straight, queer, and another sexual orientation.^9^

Descriptive statistics were calculated for the prevalence of chronic pain based on body region and widespread pain for the entire cohort. The prevalence of chronic pain by body region and widespread pain was also stratified by gender identity and sexual orientation. Logistic regression analysis evaluated whether gender identity and sexual orientation were associated with the presence of chronic widespread pain. Generalized linear models evaluated how gender identity and sexual orientation were associated with total number of body pain areas. All analyses were performed using SAS (Cary, NC).

## Results

A total of 6189 adult participants completed The PRIDE Study’s 2022 Annual Questionnaire at the time of data analysis. Among those, 792 participants did not complete pain survey items, leaving 5397 participants who completed pain survey items. A total of 2462 (45.6%) participants reported no chronic pain, leaving 2935 (54.6%) participants who reported experiencing chronic pain. The prevalence of pain by body region, widespread pain, as well results from the regression analyses are presented by gender identity group (Table 1) and sexual orientation (Table 2).

**Table 1.** Prevalence of Painful Body Regions in The PRIDE Study Participants, Stratified by Gender Identity Group and Association of Gender Identity with Total Number of Painful Body Regions and Widespread Pain.

| Body Regions (N(%)) | Cisgender Men (N=1494) | Cisgender Women (N=1825) | Non-Binary (N=1408) | Transgender Men (N=697) | Transgender Women (N=337) | Another Gender (N=225) |
| --- | --- | --- | --- | --- | --- | --- |
| Left shoulder, upper arm, lower arm | 171 (14) | 290 (19) | 336 (28.3) | 138 (24.4) | 55 (21) | 58 (31.4) |
| Right shoulder, upper arm, lower arm | 182 (14.9) | 338 (22.2) | 352 (29.6) | 166 (29.1) | 46 (17.6) | 63 (34.1) |
| Left hip/buttocks, upper leg, lower leg | 188 (15.4) | 329 (21.5) | 329 (27.7) | 150 (26.5) | 52 (19.8) | 65 (35.1) |
| Right hip/buttocks, upper leg, lower leg | 202 (16.5) | 365 (23.9) | 343 (28.9) | 152 (26.9) | 53 (20.2) | 59 (31.7) |
| Neck, upper back, lower back | 453 (33.7) | 726 (44) | 659 (52.4) | 322 (51.8) | 114 (38.1) | 107 (53.8) |
| Chest, abdomen | 85 (6.9) | 183 (11.8) | 237 (19.8) | 90 (15.7) | 33 (12.4) | 31 (16.7) |
| Right jaw, left jaw | 60 (4.9) | 183 (11.9) | 200 (16.7) | 76 (13.3) | 27 (10) | 31 (16.8) |
| Widespread pain | 211 (15.65) | 413 (24.9) | 453 (35.9) | 195 (31.3) | 64 (21.3) | 75 (37.7) |
| **Total Number of Pain Regions (Mean ± SD) (Median)** | 0.99 ± 1.46<br>0 | 1.46 ± 1.78<br>1 | 1.94 ± 2.1<br>1 | 1.75 ± 1.95<br>1 | 1.26 ± 1.6<br>1 | 2.08 ± 2.22<br>1 |
| **General Linear Model: Gender Identity (independent variable) on Total Number of Pain Regions dependent variable) (Coefficient (SE))** | -- | 0.46 (0.07)* | 0.95 (0.07)* | 0.76 (0.09)* | 0.27 (0.12)** | 1.09 (0.14)* |
| **Logistic Regression of Gender Identity as Predictors of Widespread Pain (OR (95% CI))** | -- | 1.79 (1.49, 2.15)* | 3.01 (2.5, 3.63)* | 2.45 (1.96, 3.07)* | 1.46 (1.06, 1.99)** | 3.26 (2.36, 4.5)* |

Table Legend: SD = Standard Deviation; SE = Standard Error; CI = Confidence Interval. \* = p-value <0.001; \*\* = p-value < 0.05. -- = Reference group.

**Table 2.** Prevalence of Painful Body Regions in The PRIDE Study Participants, Stratified by Sexual Orientation and Association of Sexual Orientation with Total Number of Painful Body Regions and Widespread Pain.

| Body Regions (N(%)) | Asexual/Demisexual/Gray-Ace (N=456) | Bisexual/Pansexual (N=1517) | Gay/Lesbian (N=2508) | Heterosexual/Straight (N=75) | Queer (N=1411) | Another Sexual Orientation (N=24) |
| --- | --- | --- | --- | --- | --- | --- |
| Left shoulder, upper arm, lower arm | 99 (25.7) | 251 (20) | 372 (18.1) | 18 (27.7) | 304 (25.9) | 5 (29.4) |
| Right shoulder, upper arm, lower arm | 98 (25.3) | 283 (22.6) | 404 (19.6) | 18 (27.7) | 341 (29.2) | 5 (29.4) |
| Left hip/buttocks, upper leg, lower leg | 114 (29.4) | 265 (21.1) | 404 (19.6) | 16 (24.2) | 312 (26.6) | 4 (23.5) |
| Right hip/buttocks, upper leg, lower leg | 112 (28.9) | 274 (21.9) | 431 (20.8) | 13 (20) | 341 (29.1) | 4 (23.5) |
| Neck, upper back, lower back | 198 (48.1) | 604 (44.1) | 867 (38.6) | 33 (47.8) | 671 (53.3) | 11(55) |
| Chest, abdomen | 70 (18) | 176 (13.9) | 200 (9.6) | 11 (16.7) | 198 (16.7) | 4 (23.5) |
| Right jaw, left jaw | 60 (15.4) | 156 (12.4) | 167 (8.1) | 7 (10.6) | 185 (15.7) | 2 (11.8) |
| Widespread pain (yes) | 135 (32.7) | 354 (25.8) | 478 (21.2) | 20 (28.6) | 419 (33.2) | 7 (35) |
| **Total Number of Pain Regions (Mean ± SD) (Median)** | 1.82 ± 2.12<br>1 | 1.46 ± 1.82<br>1 | 1.26 ± 1.68<br>1 | 1.86 ± 2.01<br>1 | 1.66 ± 2.01<br>1 | 1.75 ± 2.02<br>1 |
| **General Linear Model: Sexual Orientation (independent variable) on Total Number of Pain Regions dependent variable) (Coefficient (SE))** | 0.56 (0.10)* | 0.20 (0.06)** | -- | 0.39 (0.22) | 0.60 (0.06)* | 0.49 (0.41) |
| **Logistic Regression of Sexual Orientation as Predictors of Widespread Pain (OR (95% CI))** | 1.80 (1.44, 2.27)* | 1.29 (1.1, 1.51)** | -- | 1.49 (0.88, 2.52) | 1.84 (1.58, 2.15)* | 2.00 (0.79, 5.04) |

Table Legend: SD = Standard Deviation; SE = Standard Error; CI = Confidence Interval. \* = p-value <0.001; \*\* = p-value < 0.05. -- = Reference group.

## Discussion

The findings from this study highlight that regional chronic pain and widespread pain is highly prevalent in sexual and gender minority (SGM) participants in The PRIDE Study with neck, upper back, and lower back being the most common location of chronic pain. Notably, approximately one-third of non-binary and another gender persons, transgender men, and queer, asexual/demisexual/gray ace, and another sexual orientation persons experienced chronic widespread pain. Regression analyses revealed statistically significantly increased odds ratios for the presence of widespread pain in all gender identity groups with cisgender men as the reference group and in asexual/demisexual/gray ace, bisexual/pansexual, and queer sexual orientation groups with gay/lesbian as the reference group.

In a recent systematic review and meta-analysis, the overall estimated population prevalence of chronic widespread pain was 9.6%, with presumably cisgender women and men having 11.2% and 7.2%, respectively.^10^ Our findings highlight greater prevalence of widespread chronic pain in SGM persons compared to these figures, which likely are representative of a presumably cisgender heterosexual population. Our findings replicate published data in a presumably cisgender sexual minority population, which showed prevalence of widespread pain as 16.1% in gay/lesbian people, 20.1% in bisexual people, and 22.9% in persons with another sexual orientation.^3^

Our findings support the need for comprehensive studies to elucidate mechanisms for increased pain prevalence in SGM populations. Possible explanations for increased pain prevalence in SGM populations include minority stress, history of childhood or adulthood adverse life events, and limited access to affirming medical care. A limitation of the present study includes the inability to assess how these or other additional factors, including intersectionality, may contribute to chronic pain and widespread pain in SGM populations. There may be selection bias in The PRIDE Study, as SGM persons who experience pain issues may be more likely to enroll in a national SGM health study. Future studies are planned to elucidate how a variety of biopsychosocial mechanisms may influence chronic pain in SGM persons.

## Data Availability

All data produced in the present study may be available upon reasonable request to the PRIDE Study leadership team.

## Acknowledgements

The PRIDE Study is a community-engaged research project that serves and is made possible by LGBTQIA+ community involvement at multiple points in the research process, including the dissemination of findings. We acknowledge the courage and dedication of The PRIDE Study participants for sharing their stories; the careful attention of PRIDEnet Participant Advisory Committee (PAC) members for reviewing and improving every study application; and the enthusiastic engagement of PRIDEnet Ambassadors and Community Partners for bringing thoughtful perspectives as well as promoting enrollment and disseminating findings. For more information, please visit https://pridenet.org.

